# Tumour-infiltrated cortex participates in large-scale cognitive circuits

**DOI:** 10.1101/2022.12.19.22283690

**Authors:** Ayan S. Mandal, Moataz Assem, Rafael Romero-Garcia, Pedro Coelho, Alexa McDonald, Emma Woodberry, Robert C. Morris, Stephen J. Price, John Duncan, Thomas Santarius, John Suckling, Michael G. Hart, Yaara Erez

## Abstract

The extent to which tumour-infiltrated brain tissue contributes to cognitive function remains unclear. While prior studies have suggested involvement of tumour-infiltrated tissue in local circuits associated with language and motor function, it is unknown whether such tissue participates in distributed networks important for higher-order cognitive abilities like executive function. In this study, we tested the hypothesis that cortical tissue infiltrated by diffuse low-grade gliomas participates in large-scale cognitive circuits using a unique combination of intracranial electrocorticography (ECoG) and resting-state functional magnetic resonance (fMRI) imaging in four patients. We observed significant task-related high gamma (70-250 Hz) power modulations in tumour-infiltrated cortex in response to increased cognitive effort, implying preserved functionality of neoplastic tissue for complex tasks. Strikingly, we found that tumour locations corresponding to task-responsive electrodes exhibited functional connectivity patterns that significantly co-localised with canonical brain networks implicated in executive function. Finally, we discovered that tumour regions with larger task-related high gamma power elevations tended to be more functionally connected to the dorsal attention network, further demonstrating the participation of tumour-infiltrated cortex in large-scale brain networks that support executive function in health. Overall, this study contributes convergent fMRI-ECoG evidence that tumour-infiltrated cortex participates in large-scale neurocognitive circuits, reflecting preserved functionality of neoplastic brain tissue relevant to clinical management.

**Significance statement:** Gliomas interact with healthy neural circuits as they grow in the brain. Understanding these interactions is crucial for planning surgeries to remove gliomas without inducing long-term cognitive deficits. While prior studies have shown that glioma-infiltrated tissue can integrate within local functional circuits, it is unknown whether such tissue participates in large-scale whole-brain networks. Using electrocorticography, we show that glioma-infiltrated tissue responds significantly to tasks of increasing cognitive demand, reflecting its involvement in executive function processes. Using pre-operative functional neuroimaging, we found that tumour regions responsive to complex tasks were also functionally connected to large-scale networks implicated in executive function. These findings imply that gliomas participate within large-scale cognitive circuits, possibly reflecting preserved functionality relevant to clinical management.

## Introduction

Diffuse low-grade gliomas (dLGG) are slowly progressing brain tumours that infiltrate nearby healthy tissue (1). Recent evidence has indicated that gliomas integrate within their surrounding neural environment (2–5). It has been shown that glioma cells can form synapses with neurons (6) and that neuronal signalling can trigger tumour proliferation (7). Despite evidence for interference of the tumour with neural circuits (8), clinically it is known that at least to some extent glioma-infiltrated brain tissue can remain involved in motor and language functions (9), evidence that motivates the use of intraoperative mapping to preserve functionality (10). However, it is unknown whether tumour-infiltrated tissue participates in whole-brain, large-scale distributed neural circuits involved in behaviour and cognition.

To identify functionality in the surroundings of the tumour, brain tissue is typically mapped intraoperatively using direct electrical stimulation (DES), usually focusing on motor and language-related functions. However, for some functions it is more challenging to use DES, for example executive functions, a collection of cognitive processes involved in goal-directed behaviour such as attention, planning and task-switching. These functions are more complicated to map using DES because they are supported by distributed frontal and parietal regions (11–13) rather than any single, confined area. Additional information about functionality of tissue in the vicinity of the tumour may be obtained by recording brain activity directly from the surface of the brain during task performance using intracranial electrocorticography (ECoG) (8, 14). While the use of DES for mapping executive functions is scarce (15–17), we have recently employed ECoG to map peritumoural brain regions responsive to a task probing executive function (14, 18), further supported by another case report (19). Nevertheless, while DES and ECoG are useful for mapping functional regions locally, they do not provide information about the extent to which tumour-infiltrated tissue participates in distributed, whole-brain, neural circuits associated with the tested function, as can be measured with functional connectivity using fMRI data. This information is critical especially for cognitive skills such as executive functions as they are supported by coordinated activity in distributed regions in the frontal and parietal cortex (11, 12).

Assessing functional connectivity within glioma-infiltrated tissue has been traditionally controversial given that tumours are known to interfere with neurovascualar coupling, upon which the fMRI BOLD (blood-oxygenation level dependent) signal depends (20). However, it has been recently demonstrated that functional connectivity can be routinely identified within glioma-infiltrated tissue, further supporting findings from other methods like DES in establishing functionality (21, 22). While gliomas have been demonstrated to commonly localize to functional network hubs (23), it remains unclear whether glioma-infiltrated tissue participates in large-scale neural circuits important for cognitive function.

Here, we combine task-based ECoG and rs-fMRI to identify large-scale neural circuits involved in executive function in glioma-infiltrated tissue. We broadly hypothesized that tumour-infiltrated tissue would preserve its functionality and participate in large-scale brain networks. We focus on executive function for several reasons. First, executive function is very relevant to patients’ quality of life post-surgery, yet there are currently no widely implemented strategies to preserve it during glioma resection (24). Second, diffuse gliomas are most commonly observed in association cortex, and are often located in frontal brain regions associated with executive function (23). Third, we have previously validated an approach to identify a neural signature of executive function from the ECoG signal, in the form of high gamma (70-250 Hz) power that increases when task demand increases (14, 18). Finally, EF is supported by distributed whole-brain networks identified by rs-fMRI, such as the frontoparietal and dorsal and ventral attention networks (25), which we hypothesized would relate to task-based ECoG signal changes. These networks broadly overlap with task-fMRI activity patterns associated with executive function.

In this study, we tested the specific hypothesis that cortical tissue infiltrated by dLGG participates in large-scale cognitive circuits using a unique ECoG-fMRI dataset (Figure 1). We expected to find task-related power modulations in response to increasing cognitive demand in tumour-infiltrated cortex, as well as functional connectivity between task-responsive tumour locations and canonical functional networks associated with executive function.

**Figure 1.**
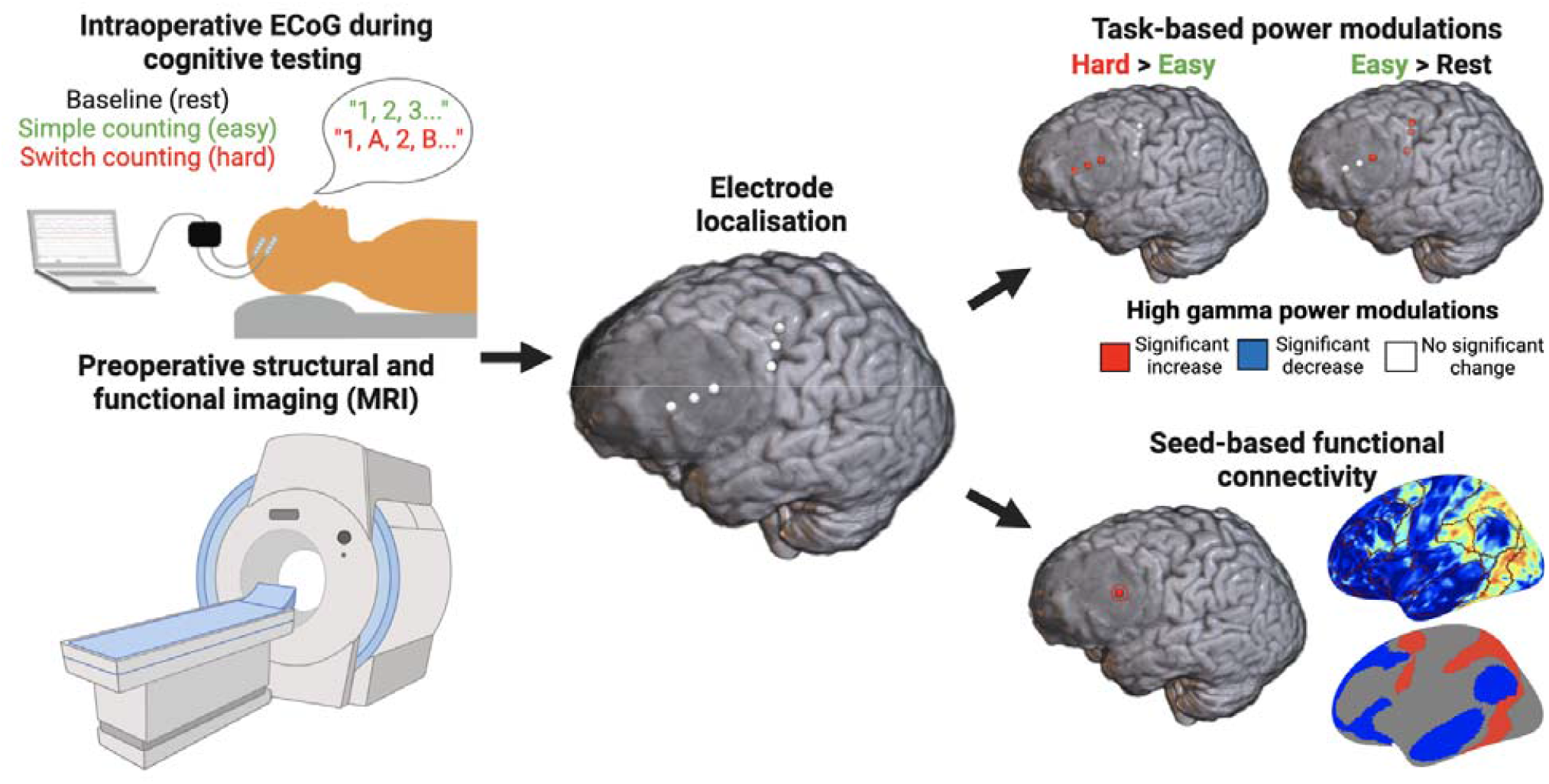
Schematic overview of the study. Participants underwent preoperative neuroimaging (bottom left) and intraoperative electrocorticography (ECoG) (top left) during three conditions (rest, simple counting, and switch counting). Locations of electrodes on tumour-infiltrated (blurred area) versus non-lesioned tissue were then localized on the preoperative structural scan (middle). Signal change in high gamma power between task conditions for each electrode was assessed (top right), reflecting task-related neural activity. Finally, whole-brain seed-based functional connectivity analyses were performed with the tumour electrode locations as seeds (bottom right), linking data across modalities. The correspondence between each resulting functional connectivity map and the topography of canonical functional networks was assessed for significance using a spin test (bottom right) (26)

## Results

### Task-related power modulations in tumour-infiltrated cortical tissue

ECoG recordings were obtained from patients intraoperatively while they were awake during three tasks with increasing cognitive demand: rest (baseline), simple counting (easy), and switch counting (hard). We calculated the percentage of signal change of high gamma (70-250 Hz) power corresponding to two task contrasts (hard > easy; easy > rest) in electrodes placed on tumour-infiltrated and non-lesioned tissue (Figure 1; Supplementary Table 1). Based on our previous findings, high gamma power increases in the hard > easy contrast were considered as reflecting areas associated with executive function (14, 18), whereas power increases in the easy > rest contrast were interpreted as reflecting cognitive processing and task activity more generally as well as speech production (27). Statistical significance of the percentage of signal changes in each electrode was assessed using a non-parametric rotation test (see Methods for details). The focus of the study is on electrodes placed on tumour-infiltrated tissue, and we show results for the electrodes that were placed on non-lesioned cortex in the same patients for reference.

Figure 2 shows each electrode rendered on each participant’s structural scan, color-coded by significance of high gamma power changes for each of the contrasts (see also Supplementary Table 1). We observed significant task-related power modulations in at least one electrode for each electrode strip placed on tumour-infiltrated tissue. For two of the three participants with frontal tumours (Patient 1 and Patient 3), significantly elevated high gamma power changes were observed for the hard>easy contrast, a measure reflecting areas involved in executive function (14). One electrode on tumour-infiltrated tissue for Patient 1 also exhibited significant high gamma increases during simple counting compared to rest, an increase that may relate to speech articulation given the proximity of this electrode to the inferior frontal gyrus (IFG). Interestingly, the other participant with a frontal tumour (Patient 2) exhibited significant high gamma power decreases for the easy>rest contrast. The participant with a tumour in the temporal lobe (Patient 4) exhibited significant high gamma increases for the easy>rest contrast, possibly reflecting speech articulation which was also localized to the superior temporal gyrus (STG) in a recent fMRI study (27).

**Figure 2.**
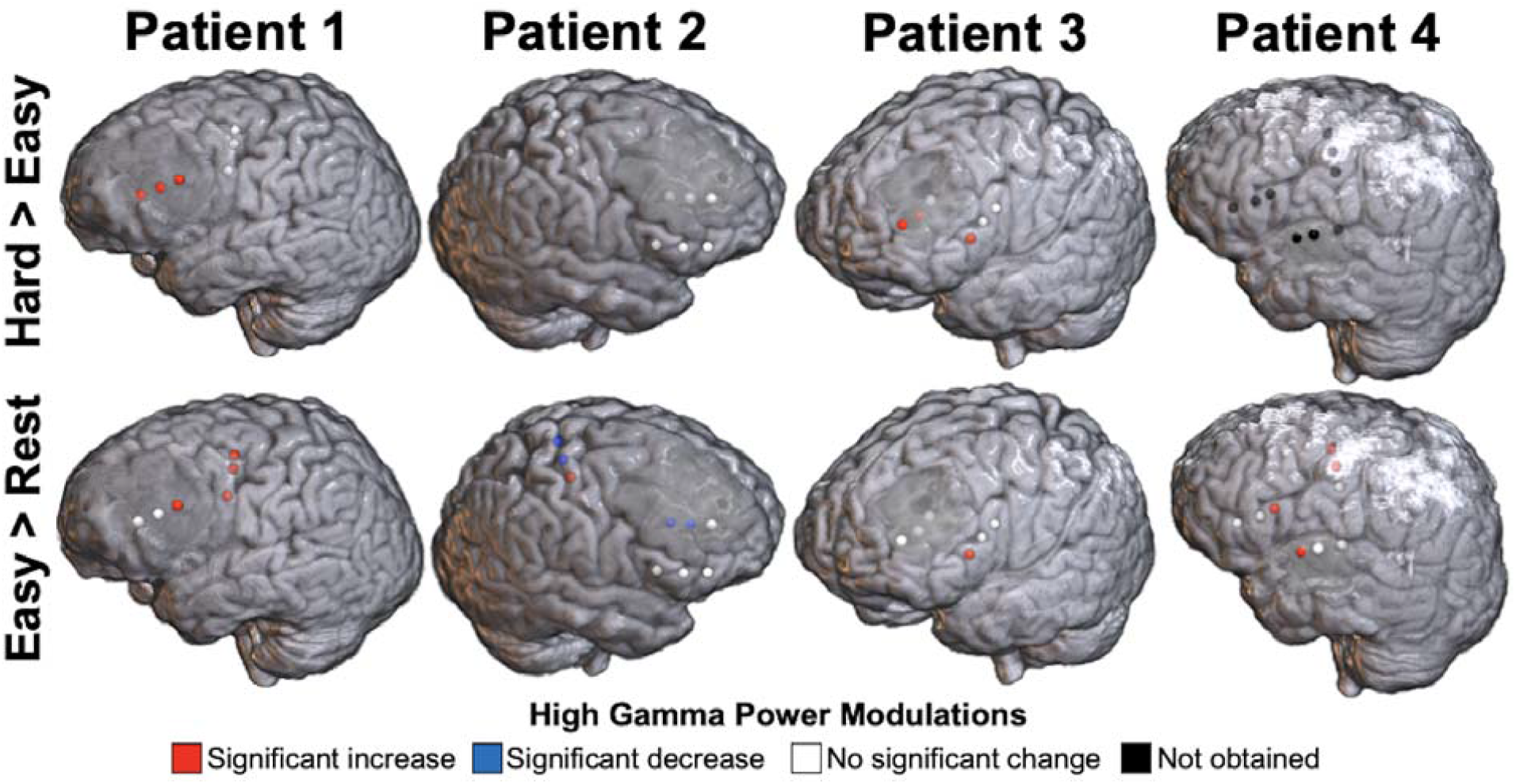
Task-related high gamma power modulations in tumour-infiltrated and non-lesioned cortical tissue. Each sphere is an electrode, colored according to the change in high gamma power for each of the two contrasts. The hard > easy contrast was not evaluated for Patient 4 because of limited performance on the switch counting task. The tumour corresponds to the blurred dark grey regions in the frontal cortex for Patients 1-3, and the superior temporal cortex for Patient 4.

### Correspondence between task-related high gamma power modulations and functional network connectivity

Next, we asked whether task-responsive tumour-infiltrated areas also participate in large-scale brain networks. Based on resting-state fMRI data of each participant, we constructed seed-based functional connectivity maps between each tumour electrode location and every voxel in the brain. We then compared these maps to canonical functional network maps derived previously from 1000 healthy individuals (28) (Figure 3A) to determine if the topology of functional connectivity from tumour-infiltrated areas retained features of healthy brain organization. Seed-based connectivity maps from the electrodes with the most elevated task-related power modulations for the hard>easy contrast for each participant (except for Patient 4, for whom the easy>rest contrast was used) are shown in Figure 3B. These maps demonstrate variations in strength of connectivity that are broadly aligned with boundaries of canonical functional networks.

**Figure 3.**
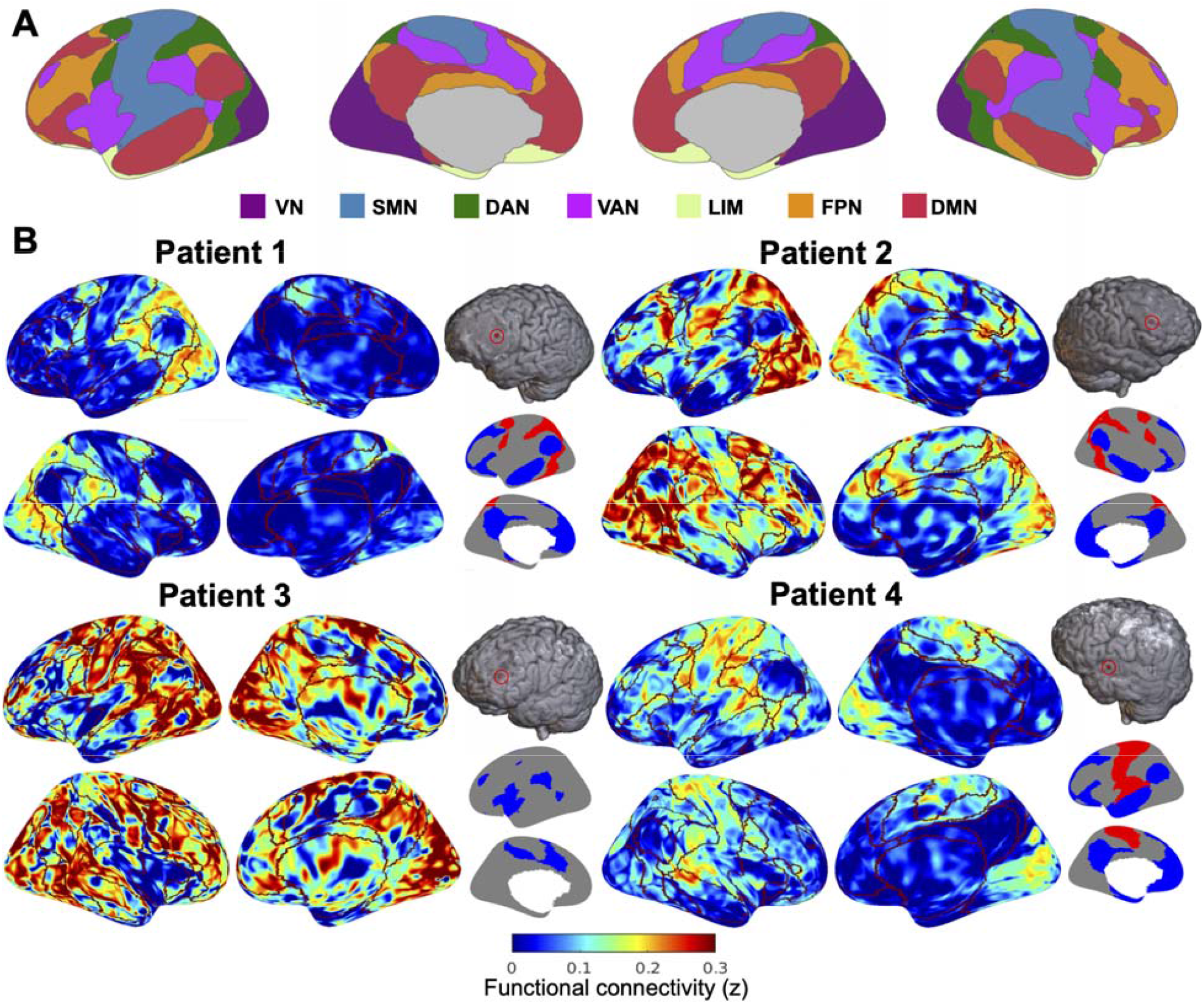
Seed-based functional connectivity of task-responsive tumour-infiltrated electrode locations. (A) Visualization of the canonical 7-network parcellation, estimated based on functional connectivity of 1000 individual (28). (B) Examples of seed-based connectivity maps from the tumour electrode location with the largest high gamma power increase for the hard>easy contrast for each participant (except for Patient 4, for whom the easy>rest contrast was used). Boundaries of the canonical networks are outlined in dark red. Seed location is shown in the top right relative to each set of surface images. The canonical networks significantly overrepresented (red) and underrepresented (blue) in the functional connectivity maps are shown on the bottom right (only one hemisphere shown for ease of visualization). VN=visual network; SMN=sensorimotor network; DAN=dorsal attention network; VAN=ventral attention network; LIM=limbic network; FPN=fronto-parietal network; DMN=default mode network.

To quantify these variations in connectivity strength, we statistically assessed the correspondence between canonical functional networks and seed-based connectivity maps using the spin test (26). The spin test statistically assesses the correspondence between two brain maps by comparing an empirical measure of their correspondence (e.g., a Pearson’s correlation) with a null distribution of the same measures when one of the maps is randomly spatially rotated (see Methods section for details). We used the spin test to determine whether certain networks were significantly over-represented (implying stronger connectivity with that network than expected by chance) or under-represented (implying weaker connectivity with that network than expected by chance) in each connectivity map. In the example task-responsive electrodes in Figure 3, for both Patient 1 and Patient 2, the dorsal attention network was significantly over-represented whereas the default mode network was significantly under-represented in the connectivity maps. The ventral attention network was significantly under-represented in the connectivity map for Patient 3. For Patient 4, the sensorimotor network was significantly over-represented whereas the default mode network was significantly under-represented. These findings demonstrate that functional connectivity from tumour-infiltrated tissue is non-random, often corresponding to canonical brain networks observed in healthy populations. Furthermore, at least in part, as demonstrated for Patients 1 and 2, over-represented networks such as the dorsal attention network correspond to the task-related activity of the electrode, both associated with attention and executive function.

Finally, we assessed the correspondence between high gamma power modulations and functional network connectivity across all participants and electrodes in tumour-infiltrated cortex. For each combination of task contrast and functional network, we performed a linear mixed effects model relating median functional connectivity to power signal changes, with Participant as the random effects variable (Figure 4). Mixed effects modelling was employed to account for repeated observations (i.e., multiple electrodes) from the same participant. A significant association was observed between dorsal attention network (DAN) connectivity and high gamma power modulations for the hard>easy contrast (χ^2^(1) = 4.31; *p* = 0.038), i.e., increased task-related high gamma power changes in tumour-infiltrated cortex were associated with increased functional connectivity of these electrodes with the DAN. No other associations between high gamma power modulations and functional network correspondence were significant (χ^2^(1) < 2.42; *p* > 0.1).

**Figure 4.**
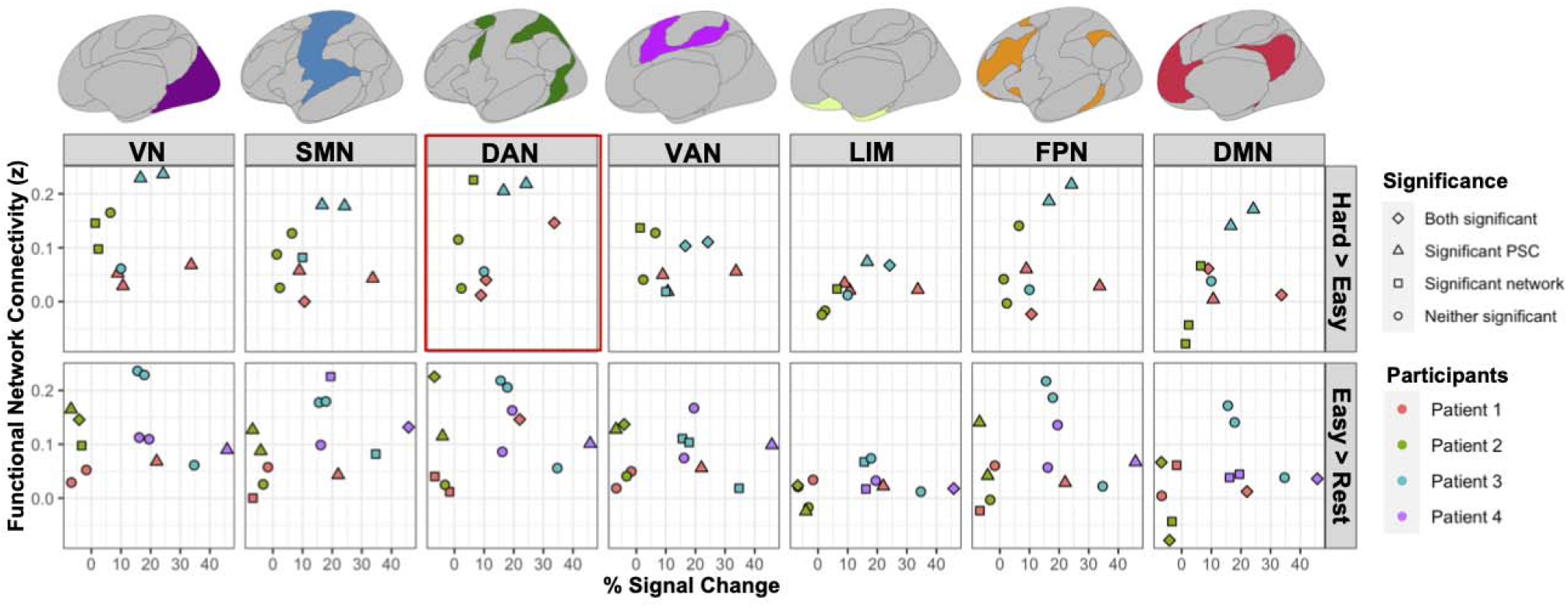
Associations between functional network connectivity and task-related power modulations in tumour-infiltrated cortex. The percentage of signal change (PSC) of each tumour electrode for each task contrast is plotted on the x-axis, against the median connectivity of that electrode location with each canonical functional network on the y-axis. Canonical functional networks are displayed above the plot with colours corresponding to the labels in Figure 3A. Datapoints are coloured by participants. Shapes denote the significance of each individual electrode’s association with the high gamma power modulation in the corresponding task contrast and network functional connectivity. Scatterplots corresponding to a significant association between network connectivity and task-based power modulations, as assessed by a mixed-effects model, are outlined in red. Note that significance of connectivity values using th spin test is determined by comparison to other functional networks associated with the same electrode. Therefore, statistical significance results may be different for similar connectivity values of different electrodes. VN=visual network; SMN=sensorimotor network; DAN=dorsal attention network; VAN=ventral attention network; LIM=limbic network; FPN=fronto-parietal network; DMN=default mode network.

## Discussion

In this study, we combined two independent modalities, ECoG and fMRI, to map regions of dLGG-infiltrated cortex involved in neurocognitive circuits associated with executive functions. Our results show convergent evidence from ECoG and fMRI that tumour-infiltrated cortex participates in large-scale cognitive networks.

Within the context of neurosurgery, glioma-infiltrated cortex is generally thought to preserve some degree of functionality, and efforts are being made to avoid the removal of critical areas and preserve functionality post-surgery (29). Evidence from both direct electrical stimulation and electrocorticography recordings have demonstrated that some tumour-infiltrated cortical regions indeed remain eloquent and functionally active (30–32) hence leading to the necessity of intraoperative mapping (29). Our ECoG findings of increased high-gamma activity with increased cognitive demand provide further support for the functionality of cortex infiltrated by dLGG and its involvement in executive function. Previous studies provided evidence for areas associated with executive function in peri-tumoural areas using DES (15–17, 19). In some cases, these areas could be detected with DES in white matter only, and not in cortical areas (15, 16). We have previously shown that high-gamma activity as recorded with ECoG from healthy tissue further away from the tumour can be used to identify areas associated with executive function. Here we further demonstrate that this neural signature of executive function is observed also in tumour-infiltrated regions, supporting the potential of using this technique for intraoperative functional mapping of executive control.

While stimulation and ECoG provide information about local areas that are necessary for these functions, little is known about whether and to what extent tumour-infiltrated areas also participate in distributed large-scale brain networks (21). This information is important because of the essential role of distributed networks in supporting function and cognition (33) and may have implications for post-surgery recovery and rehabilitation. Our findings demonstrate that task-responsive tumour-infiltrated areas, as measured with ECoG, are also part of large-scale distributed brain networks, as measured by rs-fMRI. While prior studies demonstrated that gliomas can hijack relatively local circuits involving language and motor function (34–36), our results show that tumour-infiltrated cortex participates in large-scale distributed cognitive networks important for higher-order cognition. Using ECoG, we observed activity associated with executive function in tumour-infiltrated areas. These task-related areas were functionally connected to distributed large-scale networks as measured with rs-fMRI, and in particular the DAN which has been associated with goal-directed attention, a hallmark of executive control.

In this study we show convergence of a unique combination of ECoG and rs-fMRI, linking functional data across different scales. ECoG provides information about local neurophysiological activity, independently of neurovascular coupling, thereby strengthening claims of functional persistence in glioma-infiltrated tissue. Complementarily, rs-fMRI data provide evidence for whole-brain networks that cannot be achieved with ECoG, suggesting that the functionally persistent regions of glioma-infiltrated tissue area functionally connected to distant brain networks which support executive function in health (34–36). The consistency between the results from electrocorticography and rs-fMRI provides additional validation to the latter as a non-invasive metric of tumour functionality.

A further question raised by the current research is the relationship between glioma functional integration and molecular genetic subtype. It is known that tumours of different genetic origin interact differently with the surrounding neural microenvironment. For example, it was recently shown that while cells from IDH-mutant and IDH-wildtype astrocytomas form synapses with neighbouring neurons, oligodendroglioma cells do not from such synapses (6). Direct electrical stimulation studies have also demonstrated a relationship between molecular genetic subtype and intratumoural function, as functional sites are most commonly observed in oligodendrogliomas and least commonly in glioblastomas (9). Prior literature suggests that the phenomena we report could be achieved by different mechanisms depending on the cellular composition of the tumour (6, 37). While other studies have determined that functional coupling exists between tumour voxels and the rest of the brain (21), here we demonstrate that this coupling is topologically meaningful. The finding that tumour locations exhibit functional connectivity patterns that significantly co-localize with canonical functional networks and are associated with task-responsive areas implies that BOLD signal in tumour reflects, at least in part, a meaningful signal of neural activity and is not totally confounded by neurovascular uncoupling. Our findings are consistent with recent evidence demonstrating the concordance between rs-fMRI and DES in determining tumour-infiltrated tissue involved in sensorimotor networks (22). Recent studies have also shown that intratumoural functional connectivity is prognostic of better survival and cognitive outcomes after surgery (21, 38). The relationship between intratumoural functional connectivity and relevant clinical variables (e.g., overall survival, eloquence of tissue, and cognitive outcome) suggests that rs-fMRI could be a useful, noninvasive biomarker to help guide clinical decision making (21, 38, 39). Future research should help delineate the biological significance of intratumoural functional connectivity to inform the interpretability of BOLD signal in neoplastic tissue.

Our findings may advance the understanding of the cognitive deficits associated with dLGG and the disease progression. It has been repeatedly reported that gliomas are more commonly observed in association cortex compared to sensorimotor and visual regions (23, 40, 41). Our group has previously hypothesized that the long-distance, multimodal connections characteristic of association cortex leave these regions susceptible to oncogenesis and/or tumour spread (23, 42). Our findings reported here elaborate on the interplay between glioma-infiltrated tissue and large-scale cortical networks, though it remains unclear what role these networks play in the progression of the disease. Network-based spreading of pathology along functional and structural networks has been previously reported for several other neurological conditions (43–46). Future research should address whether functional coupling between tumour-infiltrated tissue and healthy cortex could predict regions of subsequent tumour development. However, the limited sample size in our study hinders our ability to address how tumour functionality and its participation in large-scale networks varies by molecular genetic subtype or tumour location, as well as whether a similar a phenomenon may be observed in other cognitive domains.

This study provides convergent evidence from two independent modalities that cortical tissue infiltrated by dLGG participates in large-scale cognitive circuits. These findings imply functional persistence of neural circuits within glioma-infiltrated tissue, thus shedding light on mechanisms of neuroplasticity in response to neoplastic lesions. More generally, our combined ECoG-fMRI approach demonstrates the importance of cross-modality neuroimaging for advancing the understanding of functional brain networks and how they are impacted by the disease.

## Methods

### Patient recruitment

We initially recruited 21 patients suspected of a diffuse low-grade glioma and scheduled to undergo an awake craniotomy at the Department of Neurosurgery at Cambridge University Hospital. Out of these patients, five for whom an electrode strip was placed upon tumour-infiltrated cortex were included in the study. One participant was eventually excluded since electrode co-registration with MRI did not confirm placement of any of their electrodes on tumour-infiltrated cortex. Therefore, data from four participants were included and used for further analysis. Demographic and pathological information of the patients is included in Table 1. Consistent with the histological assessments, all tumours exhibited the isocitrate dehyrodgenase (IDH) mutation, pathognomonic of low-grade glioma (47). The two subjects with frontal lesions also exhibited the 1p19q codeletion, a molecular marker which distinguishes oligodendrogliomas from astrocytomas. All participants gave written informed consent to participate and were aware that the research would not affect their clinical care before, during, or after the surgery. Study protocols were approved by the East of England – Cambridge Central Research Ethics Committee (REC reference 16/EE/0151).

**Table 1.** Demographic and pathological variables for each subject. Ages are reported in ranges to remove identifying information.

| Patient ID | Age Range | Sex | Tumour location | Pathology | Molecular genetics | Notes |
| --- | --- | --- | --- | --- | --- | --- |
| Patient 1 | 40-45 | Female | Left frontal | Oligodendroglioma | IDH-mut<br>1p19q-codeletion |  |
| Patient 2 | 35-39 | Female | Right frontal | Oligodendroglioma | IDH-mut<br>1p19q-codeletion |  |
| Patient 3 | 25-29 | Male | Left frontal | Astrocytoma | IDH-mut<br>1p19q-non-codeleted |  |
| Patient 4 | 30-35 | Male | Left STG | Astrocytoma | IDH-mut | Limited performance on switch counting, therefore excluded from hard>easy contrast |

### Behavioural tasks

Patients were introduced to the tasks during standard pre-operative visits as well as a research-dedicated pre-operative assessment. Testing was performed during the surgery and following the craniotomy, just after the patient had been awakened and prior to tumour resection. ECoG recordings were obtained during a baseline condition, and during performance of two cognitive tasks. In the baseline (rest) condition, patients were asked to stay calm and remain silent for 2 to 3 minutes. The two cognitive tasks were: simple counting (1, 2, 3, up to 20; easy) and switch counting (1, A, 2, B, 3, C, up to 20; hard). These tasks were chosen to match protocols from fMRI studies probing executive function with an increased cognitive demand manipulation (11, 14, 18, 48). Each task condition was repeated for 2-5 trials based on each patient’s ability and time constraints during the surgery. Trial onset and offset markers were recorded manually on the acquisition system. One participant (Patient 4) was not able to perform the switch counting task correctly, instead uttering “1, A, 2, B, 3, B, 4, B…”. Therefore, these switch counting trials were instead labelled as simple counting, and the switch counting versus simple counting (hard > easy) contrast was not evaluated for this participant.

### MRI acquisition and pre-processing

MRI data were obtained pre-operatively using a Siemens Magnetom Prisma-fit 3 Tesla MRI scanner and 16-channel receive-only head coil (Siemens AG, Erlangen, Germany). Structural anatomic images were acquired using a T1-weighted (T1w) MPRAGE sequence (FOV 256 mm x 240 mm x 176 mm; voxel size 1 mm isotropic; TR 2300 ms; TE 2.98 ms; flip angle 9 degrees). During the same scanning session, we also acquired resting-state (eyes closed) fMRI data with a BOLD-sensitive sequence: TR=1060 ms, TE= 30ms, Acceleration factor = 4, FA = 74°, 2 mm^3^ resolution, FOV = 192×192 mm^2^, and acquisition time of 9 minutes and 10 seconds. fMRI pre-processing was performed using Independent Component Analysis (ICA) in FSL MELODIC, implemented to remove components contributing noise (49). Additionally, we performed slice timing correction, bias field correction, rigid body motion correction, grand mean scaling, and smoothing to 5-mm fixed-width half-maximum.

Masks of the tumour were drawn on the pre-operative scan using a semi-automated procedure. First, an experienced neurosurgeon (MGH) manually delineated the tumour on the pre-operative T1w image slices for each participant. Further refinement of each mask was performed using the Unified Segmentation with Lesion toolbox (https://github.com/CyclotronResearchCentre/USwithLesion) that accounts for lesion distortion by adding a subject-specific probability map before spatially warping to a reference space where tissue probability maps are predefined (50).

### Electrode localisation

The extent of craniotomy for each patient was determined purely by clinical considerations to allow for the tumour resection. Based on the craniotomy size and location, one to three electrode strips with four electrodes each were placed on the cortical surface. For the most part, electrode strips were placed in regions judged by the neurosurgeon to be healthy, but for some participants, one strip was placed on tumour-infiltrated tissue and data from these participants were included in this study. Two types of electrode strips were used, with electrode diameter either being 5 mm (MS04R-IP10X-0JH, Ad-Tech, Medical Instruments corporation, WI, USA) or 3 mm (CORTAC 2111-04-081, PMT Corporation, MN, USA). For both types, electrodes were spaced 10 mm centre to centre.

Electrode locations for Patients 2-4 were determined using an automated method with a probe linked to a stereotactic neuronavigation system (StealthStation® S7® System, Medtronic, Inc, 24 Louisville, CO, USA). For Patient 1, locations were determined using a semi-manual grid method using intraoperative photographs and a grid-like delineation of cortical sulci and gyri due to the available localisation information. Both methods are detailed below.

1. *Stereotactic neuronavigation*: Coordinates were recorded from the centre of each electrode during surgery using the neuronavigation system. The coordinates were then registered to the patient’s preoperative T1w scan. The coordinates were further shifted to correct for electrode displacements due to brain shifts following the craniotomy.
2. *The grid method:* This method is identical to the protocol described in (14) and (18), except for that the electrodes were mapped onto the participant’s T1w scan instead of the MNI template. (a) Visible major sulci were delineated on the intraoperative photographs: precentral sulcus, sylvian fissure, inferior and superior frontal sulci. Spaces between these sulci were populated by vertical lines (1.5 cm apart) to create a grid-like structure. (b) A grid was created in the same way on a 3D render of the subject’s preoperative, skull-stripped T1w scan, visualised using MRIcroMTL (https://www.nitrc.org/). (c) Voxel coordinates were manually extracted by marking the approximate locations on the grid from the 3D render corresponding to the electrode locations from the grids drawn on the intraoperative photographs.

For both methods, electrode displacements were then corrected for brain shifts by back-projecting the electrode locations onto the cortical surface along the local norm vector (51) implemented in the fieldtrip (v20160629) protocol for human intracranial data (52). Finally, electrode locations were mapped to the nearest grey matter or tumour voxel, using a nearest neighbour approach. This was done to ensure that seeds placed around the electrode locations would capture BOLD signal from cortical tissue. When a given coordinate was equidistant to two or more cortical voxels, the coordinate which ensured electrode spacing closest to 10 mm was chosen.

After electrode localisation, we constructed 2.5 mm radius spheres (to match the largest electrode diameter) around each voxel coordinate corresponding to tumour-infiltrated electrodes. We confirmed whether the electrodes were placed on the tumour-infiltrated tissue, as the surgical notes implied, by determining whether these spheres overlapped with the previously delineated tumour mask. Electrode placement on the tumour was confirmed for all electrodes from the four patients included in the study (Supplementary Figure 1).

### Electrophysiological data acquisition and analysis

Electrophysiological data were acquired using a 32-channel amplifier (Medtronic Xomed, Jacksonville, FS, USA) sampled at 10 KHz. Potential sources of electrical noise were identified and repositioned during surgery to avoid signal contamination. The data were recorded using dedicated channels on the acquisition system and two Butterworth online filters were applied: a high-pass filter at 1 Hz and a low-pass filter at 1500 Hz. A ground needle electrode was connected to the deltoid muscle. The electrodes were referenced to a mid-frontal (Fz) spiral scalp EEG electrode.

Data were analysed offline using custom MATLAB scripts and EEGLAB (v13.6.5b). The data were downsampled to 2 kHz then re-referenced using a bipolar scheme. The bipolar scheme was chosen to detect any activity changes with high spatial resolution as well as to avoid contamination of high frequency signals by scalp muscle artifacts detected by the Fz electrode. This involved re-referencing each electrode using an adjacent electrode on the same strip (i.e. 1 minus 2, 2 minus 3, 3 minus 4). As a result of the bipolar scheme, data from three electrodes from each strip were used for further analysis.

Line noise was removed by applying a notch filter at 50 Hz and its harmonics. Notch filtering was also applied at 79 Hz and its harmonics to remove additional noise observed in the data, likely due to equipment in the surgical theatre. Data were then bandpass filtered to extract activity in the high gamma (HG, 70-250 Hz) range. Instantaneous power of the timeseries was calculated by squaring the absolute amplitude envelope of the Hilbert transformed data. We focused on high gamma power given prior evidence of the concordance of this frequency band with BOLD signal and neuronal activity associated with cognitive processing (53), as well as evidence linking increase in HG activity following increased cognitive demand to executive function (14, 18).

The power timeseries data were then segmented into the appropriate conditions and trials. Because trial onset and offset markers were manually recorded, two seconds from the beginning and end of the rest trial and one second from each task trial were excluded to account for error related to human reaction time. For the switch counting trials, a further three seconds were excluded from the beginning of each trial to discard the initial easy phase of this task (1, A, 2, B, 3, C).

To link neural activity to certain cognitive processes, we calculated the percentage of signal change in high gamma power for two task contrasts: hard>easy and easy>rest. Based on our previous findings (14, 18), increases in HG activity in the hard>easy contrast indicate regions associated with executive function, whereas for the easy>rest contrast these changes serve as indication for task-responsive regions more generally as well as regions involved in speech production. We computed a percentage of signal change for each contrast by first calculating the average power for each condition across all trials and time points. For each contrast (hard>easy, easy>rest), the percentage of signal change was then computed as: [(power in condition 1/power in condition 2) – 1] * 100.

A permutation testing approach was used to statistically test for power modulations in each electrode while controlling for temporal autocorrelation of the signal (14). For each electrode and contrast, the instantaneous power timeseries of all task trials from both conditions in the contrast were concatenated serially to form a loop. To close the loop, the end of the last trial was joined to the beginning of the first trial. All trial onset and offset markers were then shifted using the same randomly generated jitter, allowing the power timeseries values to “rotate” along the data loop. This rotation approach was used to generate surrogate power data while preserving trial lengths and the temporal autocorrelations in the data. After the rotation, the mean power (for each condition) and percentage of signal changes (for the conditions in the contrast) was computed based on the new trial markers. By applying this rotation approach to the power timeseries as opposed to the raw signal, we ensured that no artefacts in the form of sudden power changes at the points of trials concatenation were introduced. This procedure was repeated 100,000 times with a different random jitter for each permutation to create a surrogate distribution against which two-tailed statistical significance (alpha = 0.05) was calculated for each contrast within each electrode. Significance testing was performed for both tumour-infiltrated and healthy-appearing electrodes to provide context for task-related power modulations in tumour-infiltrated cortex.

### Seed-based functional connectivity analysis

For each electrode placed on tumour, we conducted seed-based functional connectivity analysis to determine whether the corresponding cortical tissue was functionally coupled to canonical resting state networks. Using FSL software (49), the previously mentioned 2.5 mm radius spheres placed around each tumour electrode coordinate were first masked such that they covered only brain parenchyma, then mapped into the space of each participant’s functional scan. Next, the mean BOLD timeseries of voxels within the seed was calculated, then cross-correlated (Pearson’s) with the BOLD timeseries of each other brain voxel. A Fisher’s R to Z transformation was performed on the resulting map, which was then smoothed at 5mm full-width half maximum. Finally, the seed-based functional connectivity map was mapped back onto the space of each participant’s native structural scan.

The volumetric seed-based functional connectivity maps were then projected onto surface vertices for the purposes of visualisation and statistical inference of canonical network correspondence. Using Freesurfer reconstructions of each participant (54), as well as Connectome Workbench code (55), each connectivity map was mapped onto fsaverage (∼164k vertices) using nonlinear surface-based registration. Each map was visualised with the boundaries of the canonical Yeo 7-network parcellation outlined on the surface (21). This canonical 7-network parcellation was constructed previously by employing a clustering approach to rs-fMRI data from 1000 college-aged individuals (28).

To determine whether the seed-based functional connectivity maps significantly co-localised with specific canonical functional networks, we conducted the spin test (26). This approach conducts a nonparametric permutation test to infer statistical significance while controlling for spatial autocorrelation. First, the empirical median connectivity values were calculated within each canonical functional network. These values were evaluated for significance by comparing to a distribution of analogous Z values computed from randomly-rotated surrogate maps. The surrogate maps were generated by projecting the original map onto the fsaverage sphere, randomly rotating the sphere, then projecting back onto the pial surface. Equivalent rotations were performed for each hemisphere, such that bilateral symmetry was preserved for each surrogate map. Values from the medial wall land on the cortical surface during these rotations were excluded from the analysis. Median connectivity values were then computed for each canonical network in the surrogate data. We generated 10,000 surrogate maps for each seed-based functional connectivity map. The percentile of median empirical connectivity within each network compared to the null distribution of each network’s median connectivity values was then used to estimate two-tailed *P*-values (alpha = 0.05), indicating which networks are significantly over-represented or under-represented in each connectivity map (i.e., connected to the seed more/less than expected by chance).

### Associations between task-related power modulations and functional network correspondence

Finally, we were interested in the relationship between task-based power modulations derived from ECoG with the functional connectivity maps derived from rs-fMRI in tumour-infiltrated cortex. Using the *lme4* package in R, we performed 14 (2 ECoG contrasts x 7 fMRI networks) linear mixed effects models relating median functional connectivity within each canonical network with percentage of signal changes from each task contrast. Percentage of signal change was the response variable, while functional connectivity and participant ID were modelled as the fixed and random effects, respectively. Statistical significance of the resulting mixed effects models was evaluated using a Type II Wald chi-square test computed using the *car* package in R.

### Visualisation software

The schematic diagram in Figure 1 was created using BioRender (biorender.com). Visualisation of electrode locations on each patient’s preoperative scan was done using 3D rendering in MRIcroMTL (https://www.nitrc.org/). Surface maps were constructed using code adapted from The Computational Brain Imaging Group head by Thomas Yeo (https://github.com/ThomasYeoLab/CBIG). Brain plots of the Yeo networks in Figure 3A and Figure 4 were created using ggseg in R (https://github.com/ggseg/ggseg).

## Data Availability

All data produced in the present study are available upon reasonable request to the authors.

