## Supplementary Material for "Tumour-infiltrated cortex participates in large-scale cognitive circuits"

**SUPPLEMENTARY MATERIALS**

| **Participant** | **Strip** | | **Channel** | **Tumour?** | **Hard > Easy**  **(% Signal Change)** | **Easy > Rest**  **(% Signal Change)** |
| --- | --- | --- | --- | --- | --- | --- |
| Patient 1 | S1 | EL1 | | Yes | **10.7** | -6.8 |
|  | S1 | EL2 | | Yes | **8.9** | -1.7 |
|  | S1 | EL3 | | Yes | **33.7** | **22.0** |
|  | S2 | EL1 | | No | -67.6 | **371.5** |
|  | S2 | EL2 | | No | -81.2 | **732.3** |
|  | S2 | EL3 | | No | -86.5 | **1033.2** |
| Patient 2 | S1 | EL1 | | No | 2.0 | -9.9 |
|  | S1 | EL2 | | No | -2.3 | -13.5 |
|  | S1 | EL3 | | No | -4.0 | -4.8 |
|  | S2 | EL1 | | Yes | 2.4 | -3.3 |
|  | S2 | EL2 | | Yes | 1.3 | **-4.1** |
|  | S2 | EL3 | | Yes | 6.4 | **-6.9** |
|  | S3 | EL1 | | No | -5.9 | **27.3** |
|  | S3 | EL2 | | No | 2.5 | **-22.5** |
|  | S3 | EL3 | | No | 8.1 | **-27.7** |
| Patient 3 | S1 | EL1 | | No | **20.1** | **16.4** |
|  | S1 | EL2 | | No | 7.6 | -4.3 |
|  | S1 | EL3 | | No | 14.8 | 18.9 |
|  | S2 | EL1 | | Yes | **24.2** | 15.6 |
|  | S2 | EL2 | | Yes | **16.6** | 17.9 |
|  | S2 | EL3 | | Yes | 10.0 | 34.7 |
| Patient 4 | S1 | EL1 | | Yes | - | **45.8** |
|  | S1 | EL2 | | Yes | - | 16.1 |
|  | S1 | EL3 | | Yes | - | 19.5 |
|  | S2 | EL1 | | No | - | 1.7 |
|  | S2 | EL2 | | No | - | 6.6 |
|  | S2 | EL3 | | No | - | **10.1** |
|  | S3 | EL1 | | No | - | **70.1** |
|  | S3 | EL2 | | No | - | **78.7** |
|  | S3 | EL3 | | No | - | 41.1 |

**Supplementary Table 1. High gamma power modulations in electrodes placed on lesioned and non-lesioned cortex.** Statistically significant power modulations are in bold (*p* < 0.05). Strips are numbered such that the most lateral non-motor cortex strips are first, and the motor cortex strip is always last. Electrodes in the non-motor cortex strips are numbered from anterior to posterior, and the motor cortex strip is numbered from lateral to medial.

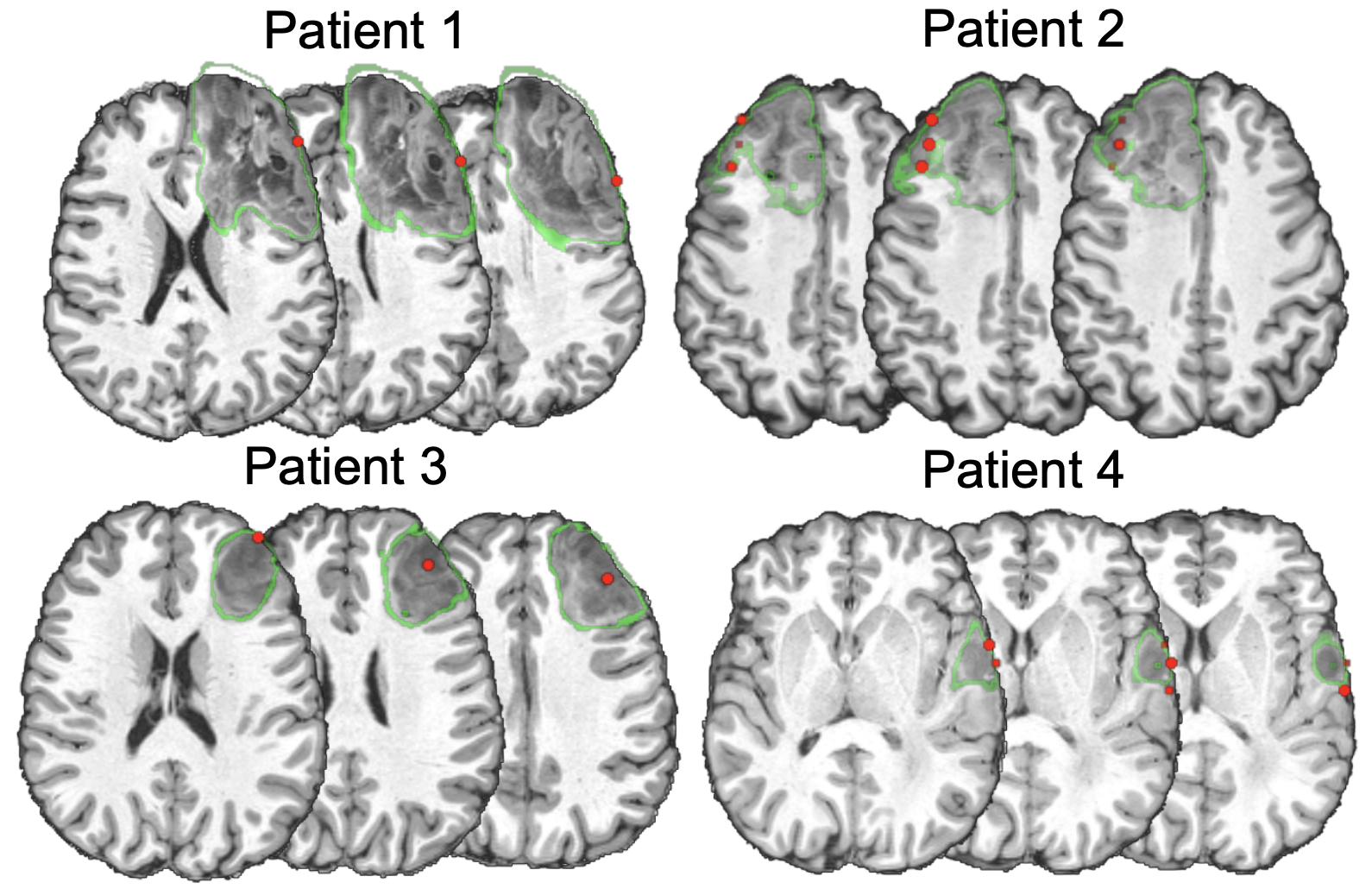

**Supplementary Figure 1. Placement of electrodes within tumour margins.** Electrodes placed on tumour-infiltrated cortex are shown in red, while the margins of the tumour for each patient are drawn in green.
